# Closed-Loop, Subgaleal Intersectional Short-Pulse Stimulation for the Treatment of Therapy-Resistant Epilepsy in Adults

**DOI:** 10.1101/2025.09.27.25334859

**Authors:** Zoltán Chadaide, Dániel Fabó, Miklos Szoboszlay, Lívia Barcsai, Andrea Pejin, Bálint Horváth, Márton Görög, Tamás Földi, Lili Ambrúzs, Tamás Laszlovszky, László Halász, Márton Huszár-Kis, Nóra Forgó, Gábor Szilágyi, Áron Négyessy, Anna Kelemen, Zsófia Jordán, Ákos Ujvári, Anna Sákovics, Anita Kamondi, György Buzsáki, Orrin Devinsky, Loránd Erőss, Antal Berényi

## Abstract

One-third of epilepsy patients do not fully respond to antiseizure medications (ASM), are not candidates for curative surgical interventions, or have unsuccessful surgical therapies. Safe and effective therapies remain limited for these treatment-resistant epilepsy (TRE) patients. Several palliative neurostimulation devices were approved by the US Food and Drug Administration (FDA) and other regulatory agencies: vagus nerve stimulation (VNS), responsive neurostimulation (RNS), and deep brain stimulation (DBS). Only RNS is responsive and can continuously monitor electroencephalographic (EEG) activity, but its closed-loop stimulation is limited by spatiotemporal accuracy. VNS, RNS, and DBS require invasive implantations that can cause adverse outcomes. We developed a minimally invasive tool with automatic seizure detection that can deliver therapeutic intersectional short-pulse (ISP) stimulation to terminate pathological brain activity. The therapy exerts an immediate effect, reducing seizures from the onset of treatment, without the prolonged adaptation period typically required by other neurostimulation approaches. We assessed the safety, feasibility, and effectiveness of ISP stimulation delivered transcranially through subgaleal electrodes in epilepsy patients with focal seizure and with Lennox-Gastaut syndrome. Ictal ISP stimulation reduced seizure duration by 52% in average, modulated spectral content, and inhibited secondary generalization. Among patients with multiple daily seizures, closed-loop ISP stimulation reduced seizure incidence >80% within days of inpatient treatment. Minimally invasive implantation strategy with precise, closed-loop ISP delivery can safely and effectively reduce seizure activity in TRE patients. This trial is registered at ClinicalTrials.gov, identifier NCT07041619.

## Introduction

A third of epilepsy patients suffer ongoing seizures despite available anti-seizure medications (ASMs), resective surgery and neurostimulation devices (treatment-resistant epilepsy).^1,2^. While surgical interventions can be curative, fewer than 50% of TRE patients are surgical candidates, with seizure control in <60% and risks of permanent neurological deficits^3,4^. Neuromodulation offers a promising alternative for refractory clinical populations unresponsive to conventional therapeutic modalities.

Three neurostimulation devices are approved by the US Food and Drug Administration (FDA) for open-loop and responsive (i.e. on-demand) stimulations. Anterior thalamic nucleus deep brain stimulation (ANT-DBS) and vagus nerve stimulation (VNS) apply open-loop stimulation. Both approaches decrease cortical excitability and desynchronize the epileptic network, reducing pathological synchrony^5–7^. ANT-DBS reduces θ and α band synchronization on EEG^8^, while VNS reduces θ band spectral power and desynchronizes EEG activity in responders^9,10^. Responsive neurostimulation (RNS) applies closed-loop stimulation directly to epileptogenic zones, disrupting pathological oscillations during seizure transitions^11,12^. RNS requires invasive implantation under the skull and can only target two seizure zones^13^. DBS and RNS have higher rates of intracerebral bleeding and are not clearly more effective than VNS^14^.

To address these limitations, we developed a subgaleal electrode-based system combined with the novel intersectional short-pulse (ISP) stimulation to enable minimally-invasive, high-intensity neuromodulation (Extended Data Fig. 1). ISP consists of ultra-brief, distributed pulses to maximize electric field strength in target areas while minimizing adverse effects on non-target tissues^15,16^. Early preclinical studies demonstrate its efficacy in disrupting pathological oscillations and reducing seizures in animal models^16–18^. These advancements pave the way for clinical applications using closed-loop ISP stimulation in refractory epilepsy patients.

## Results

We enrolled 15 patients from November 2021 to September 2025 to investigate ISP stimulation’s feasibility, safety, and efficacy to suppress seizures. Two patients withdrew consent before implantation. In the remaining thirteen patients, 2283 hours of video-EEG data were recorded during 127 patient-days during inpatient monitoring. One patient’s measurement was terminated early due to cable breakage, while twelve completed the experimental protocol (Extended Data Fig. 2). Another patient was excluded from the analysis due to a lack of identifiable ictal EEG waveforms. A detailed description of the seizure onset zone (SOZ) localization, ISP sequence planning, subgaleal electrode positioning, and post-operative validation can be found in Extended Data Fig. 3.

We observed 50 Grade 1 and 26 Grade 2 adverse events (AEs) throughout the 127 experimental days for 13 patients. In addition, two Grade 3 AEs occurred (i.e. psychosis related to antiseizure medication tapering and fever due to urinary infection), both unrelated to the subgaleal implantation procedure or ISP stimulation. No serious AEs were observed in relation to the therapy, and no Grade #4 or #5 AEs occurred. All AEs fully resolved, spontaneously (N = 38) or after therapy (N = 40). The summary of the observed AEs is provided in Extended Data Table 1.

To ensure patient comfort, individual ISP stimulus intensity thresholds were determined using a Visual Analog Scale (VAS). Perception limits (8.1 ± 3.9 mA) and tolerability thresholds (28.2 ± 11.2 mA) varied across patients (N = 13), with stimulus intensity strongly correlating with VAS scores (r^2^ = 0.81, 303 observations; Fig. 1a–c).

**Fig. 1.**
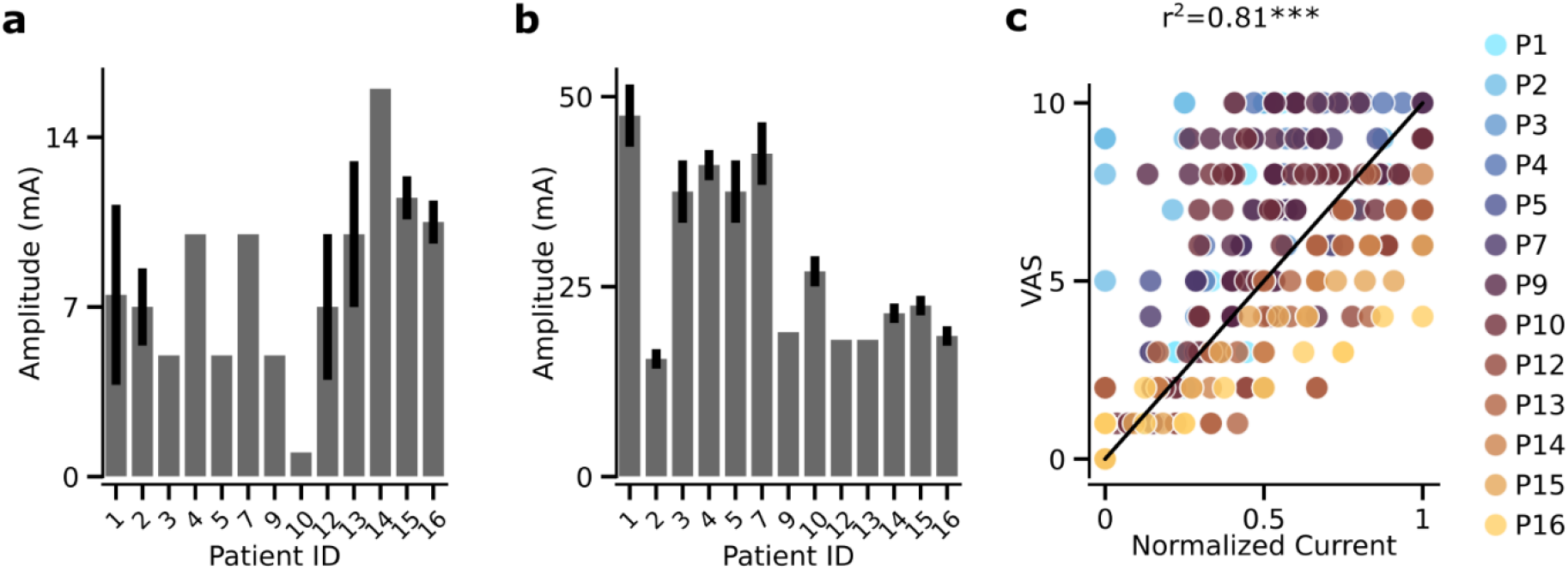
ISP stimulation tolerance of patients. **a**, Perception limits of ISP stimulation across individual patients are highly variable. **b**, Highest stimulation intensities of the patients show pronounced individual variability. **c**, Subjective VAS values of the patients are highly correlated with the stimulation intensities (r^2^ = 0.81, ***p<0.001). Measurements below perception threshold are discarded and current values between perception threshold and maximum tolerable level are normalized to [0 – 1] for each patient.

ISP stimulation aimed to reduce seizure duration, severity, and secondary generalization. The experimental paradigm was structured in two phases: an initial monitoring period to capture patients’ habitual seizures followed by a treatment period with closed-loop ictal ISP stimulation. We recorded 273 control and 231 stimulated seizures. A single dose ISP stimulation consisted of three repeats of 50-ms-long half-sine modulated ISP pulse trains. The interstimulus interval within the trains of the three repeats were adjusted to the prominent seizure frequency to stimulate the same phase of the subsequent oscillation cycles (Extended Data Fig. 1). A representative example from Patient 2 with a control and a stimulated epileptic seizure is shown in Fig. 2a and on Extended Data Video 1.

**Fig. 2.**
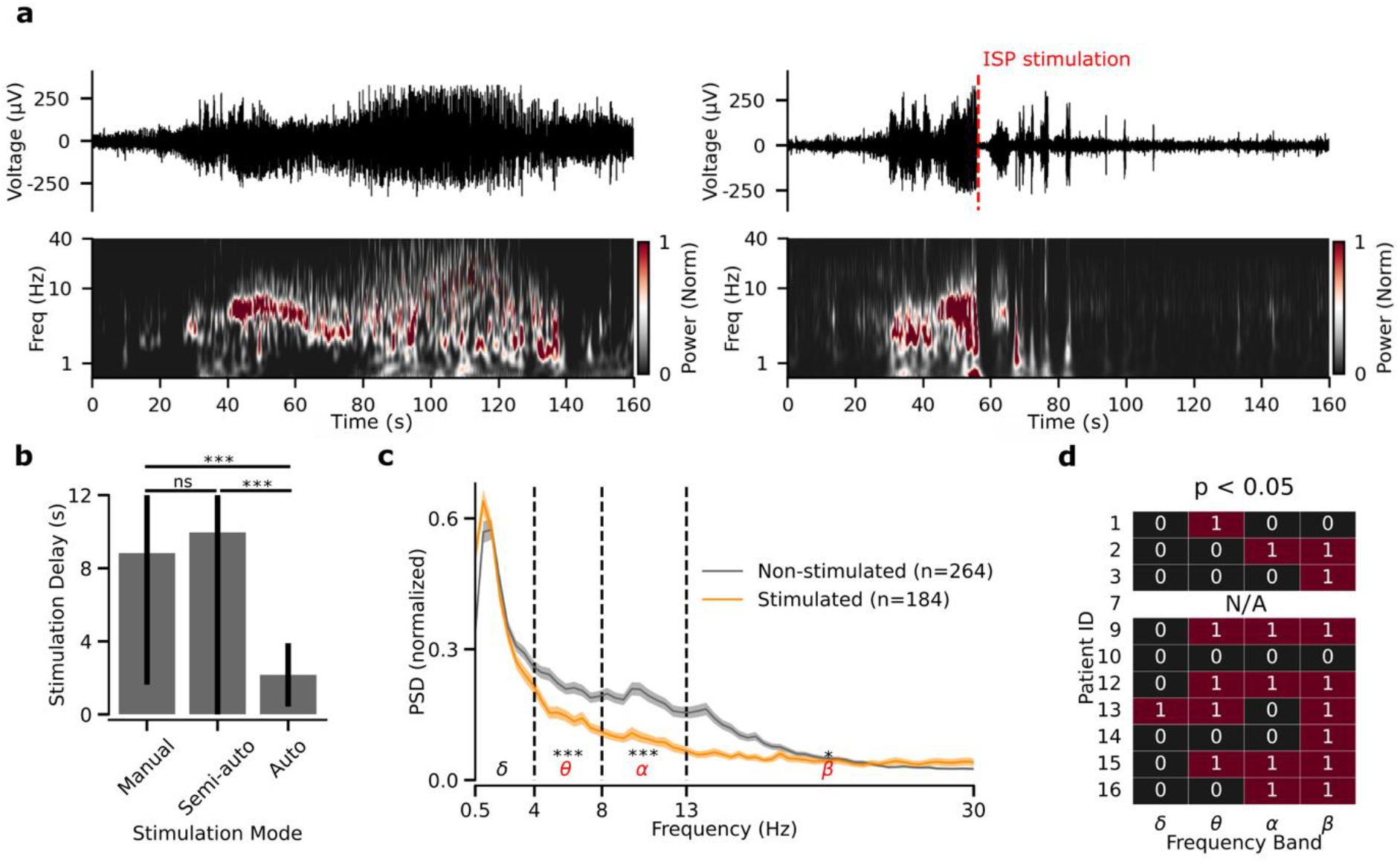
ISP stimulation modifies the spectral content of epileptic seizures. **a**, Representative examples of a control, non-stimulated (left) and a stimulated (right) epileptic seizure from Patient #2. Top: Filtered single channel EEG trace, bottom: wavelet spectrogram of the EEG trace above. Dashed red line on the right shows the time of the ISP stimulation and its immediate effect on seizure termination. **b**, Mean stimulation delay of the three stimulation modes used during the study. While there was no difference between the manual and semi-automatic modes, fully automatic stimulation delays were significantly shorter than both other modes (Kruskal-Wallis test with Dunn’s post hoc test and Bonferroni correction; ***p<0.001). **c**, Mean normalized PSD of control, non-stimulated (gray), and stimulated (orange) epileptic seizures (mean ± s.e.m.). At the population level, the spectral content of the ISP-stimulated seizures significantly decreased in the *θ, α* and *β* frequency bands (two-sample Kolmogorov-Smirnov test, *p<0.05, ***p<0.001). **d**, ISP stimulation’s effect on the epileptic seizures’ spectral content is variable at the per-patient level. Red squares indicate the frequency bands where the ISP stimulation significantly changed the spectral content of the stimulated seizures compared to the control, non-stimulated ones (two-sample Kolmogorov-Smirnov test). The control seizure of Patient #7 was omitted from the analysis due to corrupted EEG signals.

Stimulation trigger was aided by an online machine learning algorithm (Extended Data Fig. 4). Three stimulation modes were available on our custom SeizureStop device: “manual” (in case the detector algorithm missed the seizure), “semi-automatic” (the detector algorithm signaled at the seizure onset, and an expert’s confirmation was required for stimulation), and in high-confidence cases, an “automatic”, closed-loop stimulation mode was enabled. The mode-dependent stimulation delays were similar in case of the first two. However, the automatic stimulation had a significantly shorter delay after seizure initiation (Fig. 2b).

After the first ISP was delivered, ictal patterns and semiology were evaluated. If the seizure persisted, the stimulation was repeated up to five times. Most seizures were stimulated once or twice.

ISP stimulation altered the spectral content of the EEG in eight out of nine patients. At the population level, it decreased the power of the θ, α and β frequency band during seizures, similarly to ANT-DBS (Fig. 2c). We observed a substantial heterogeneity of ISP stimulation’s effect at the individual patient’s level (Fig. 2d).

The stimulation abruptly ceased the ictal electrographic patterns in the EEG and consequently shortened the seizure durations in 10/11 patients (Fig. 3a). In the eight patients with high seizure frequencies, this difference was statistically significant; the same trend was observed for the other two patients. One patient’s seizure durations were unaffected by ISP stimulation. Overall, ISP-stimulated seizures were 52 ± 35% shorter than controls, even with the substantial delay introduced by manual and semi-automatic stimulation modes. After compensating for the delays introduced by the supervised enabling of stimulation, seizure shortening was 66 ± 39% (Fig. 3b; p < 0.001; N = 272 baseline vs. 224 stimulated seizures from 11 patients). Comparing the per-patient mean differences yielded 50 ± 25% raw shortening and 61 ± 30% delay-compensated shortening, respectively (p = 0.005; N = 11 vs. 11 mean values, paired t-test; for per patient statistical details, see Extended Data Table 2.).

**Fig. 3.**
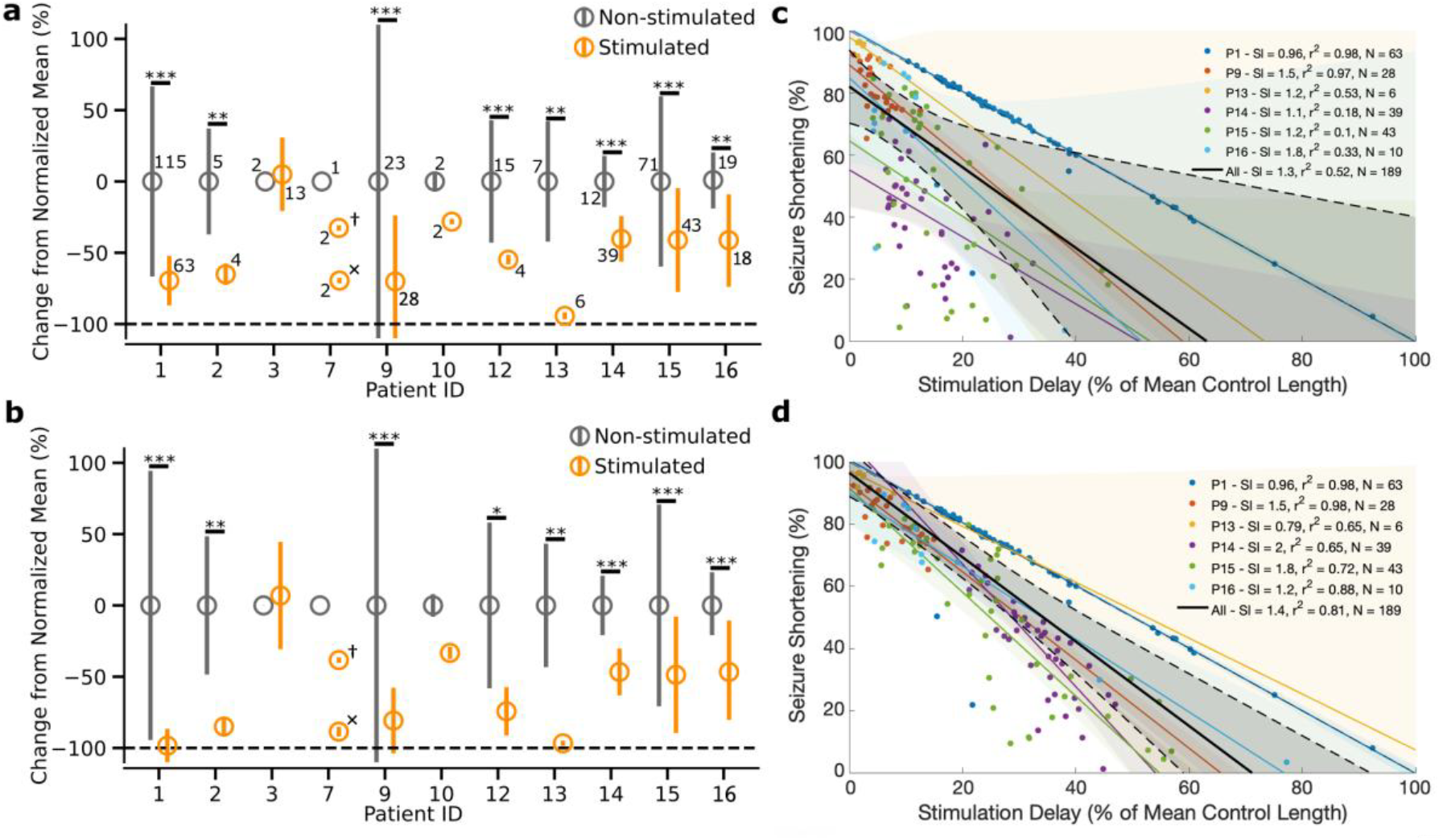
ISP stimulation efficiently terminates epileptic seizures. **a**, Epileptic seizure length of control, and stimulated seizures with or without clinical correlates. Where the number of observations was sufficiently high (i.e., n >= 4), a two-sample Kolmogorov-Smirnov test determined statistically significant shortening of seizure durations evoked by ISP stimulation (**p<0.01, ***p<0.001). Other patients showed a similar trend. Numbers indicate the sample sizes; horizontal dashed line represents immediate seizure termination. For Patient 7, ^†^ and ^×^ denote left frontal and bilateral ISP stimulation paradigms, respectively. **b**, Same as **a**, but durations of the stimulated seizures were calculated after the first ISP stimulation, and the control seizures’ length were compensated by the average delay of the first ISP stimulations in each patient (*p<0.05, **p<0.01, ***p<0.001). Note that seizures of Patients 1, 2, 9 and 13 almost immediately terminated after the first ISP stimulation. **c**, Seizure shortening inversely correlates with the temporal delay of the first ISP stimulation (***p<0.001). **d**, Same as **c**, but in case of multiple ISP stimuli delivered, their mean was calculated and plotted against seizure shortening (***p<0.001 for all patients in both comparisons. Seizure length and stimulation delays were normalized to the mean duration of the control seizures in each patient, diagonal depicts immediate seizure termination at stimulus delivery. Solid lines: linear regression, shaded area: 95% confidence interval from n = 1000 bootstraps, Sl: slope of the regression lines) Note, that patients with less than ten stimulated seizures and seizures with incomplete stimulation were excluded from this analysis.

Seizure shortening was inversely correlated with the temporal delay of the first ISP stimulation relative to seizure onset. Delayed stimulation made the seizures 1.29 ± 0.32× longer compared to the delay of the stimulation onset (r^2^ = 0.52; p <0.001; N = 189 seizures from five patients; Fig. 3c). For multiple ISP stimuli delivered within a single epileptic seizure, seizures durations became 1.4 ± 0.5× longer than the increase in the mean temporal delay of the stimulation (r^2^ = 0.81; p < 0.001; N = 189 seizures from five patients; Fig. 3d). In the latter case, measured seizure duration was further biased by considerable delays introduced during manual reassessment of seizure persistence. Enabling an automated stimulation repeat protocol could potentially further reduce seizure duration. These results suggest that targeting the early onset of the seizures may result in more effective seizure termination.

ISP stimulation effectively prevented secondary generalization in two of the three patients with secondary seizure generalization (Fig. 4a). For Patient 7, limited efficacy of the initially administered ISP sequence targeting the left frontal lobe necessitated a shift to a bilateral stimulation protocol. This adjustment led to significantly improved seizure abbreviation and effectively prevented generalization. The third patient (Patient 10) experienced rapid generalization of seizures within less than 5 seconds. Stimulation was only applied during the generalized phase, as semiautomatic mode necessitated manual confirmation, resulting in stimulation delays of 9 and 12 seconds.

**Fig. 4.**
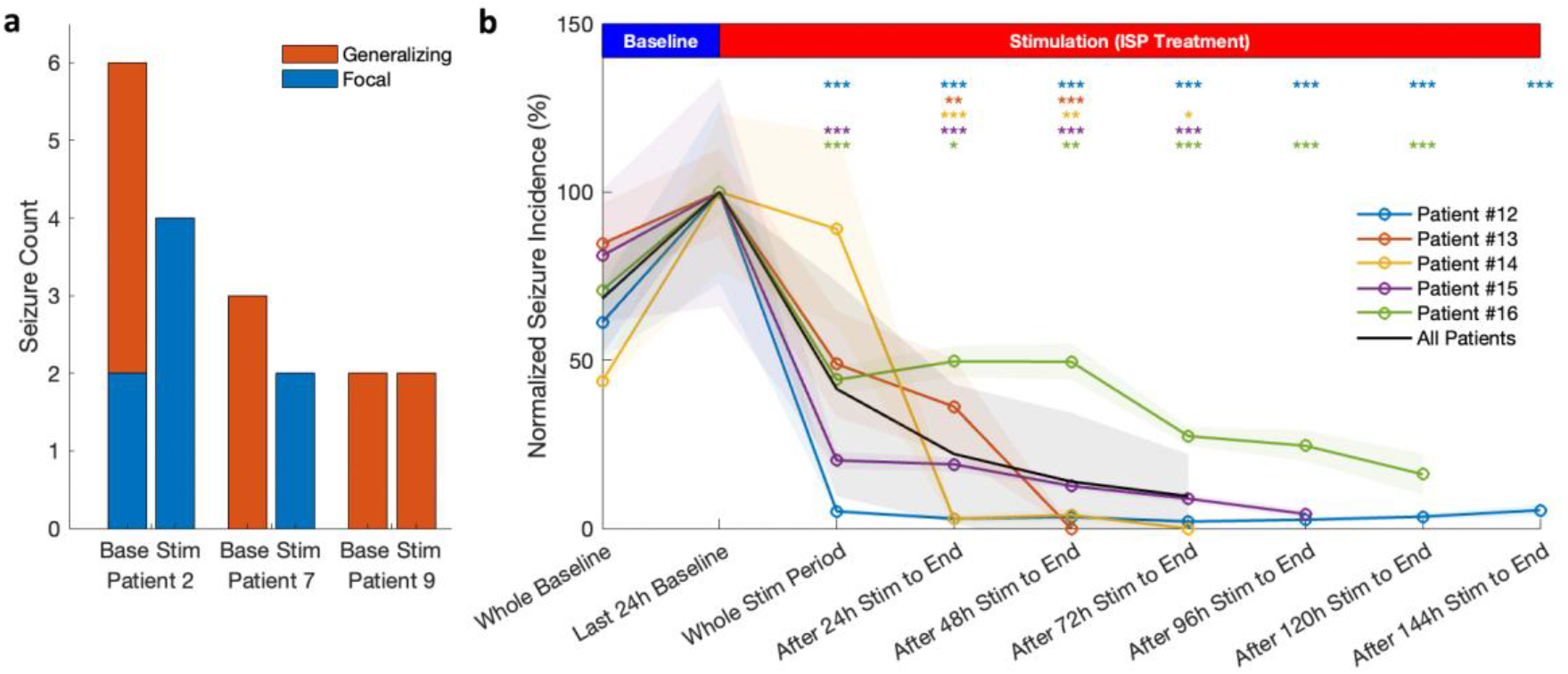
ISP stimulation efficiently prevents generalization and decreases seizure incidence. **a**. Number of generalized seizures in patients susceptible to secondary generalization. ‘Base’ and ‘Stim’ refer to baseline and stimulated seizures, respectively. For Patient 7, note that seizures stimulated with the ineffective left temporal ISP sequence are categorized within the baseline group. **b**, Figure shows the electroclinical seizure incidence rates with normalized to the baseline incidence rate from the appearance of the last 24 hours before the beginning of ictal ISP stimulation. To reveal the gradual build-up of the therapeutic effect, each individual data point shows the incidence rates during the remaining period of the experiment after being exposed to a certain amount of ISP stimulation. For the individual patients, mean ± s.e.m. is shown for better visibility. Population data shows the average of the individual patients’ results and their standard deviation. Asterisks denote the significance levels against the baseline incidence rate (for detailed analysis protocol, per-patient numerical data, see Extended Data Fig. 5).

Based on the favorable safety profile observed in the first cohort, we selected a second cohort of patients with relatively high baseline seizure incidence and performed continuous, high-coverage closed-loop stimulation of epileptiform patterns. In this cohort, seizure frequency was assessed using progressive cumulative incidence analysis to evaluate potential effects on seizure incidence. For the stimulated period, incidence was recalculated cumulatively from different starting points (entire stimulation window, 24 h after onset, 48 h after onset, etc.), thereby revealing the gradual build-up of the therapeutic effect.

Closed-loop ictal ISP stimulation progressively reduced seizure incidence (i.e. both electrographic and clinical correlates) in all five patients, with reductions evident within just a few days of stimulation (Fig. 4b, Δ_Incidence_ = -55.8 ± 42.4 %, -80.9 ± 36.7 %, and -96.0 ± 19.3 %, after 24, 48 and 72 hours of treatment, respectively, median ± IQR; N = 5 patients). In three patients with a high baseline seizure burden, ISP stimulation achieved complete seizure freedom for at least one full day prior to explantation. In the remaining two patients, seizure incidence was markedly reduced with a clear trend toward seizure freedom. Patient-level seizure counts and incidence values are provided in Extended Data Fig. 5. A similar progressive decrease was also observed in the total time spent in seizures (Extended Data Fig. 6, Δ_TiS_ = -89.7 ± 34.9 %, -98.2 ± 14.9 %, and -97.5 ± 12.5 %, after 24, 48 and 72 hours of treatment, respectively, median ± IQR; N = 5 patients).

## Discussion

This first-in-human trial of ISP stimulation delivered via subgaleal strip electrodes demonstrates promising safety and efficacy for TRE patients. ISP stimulation was tailored to individual patients’ ictal patterns, demonstrating reduced seizure duration and spread, as well as reduced seizure incidence compared to unstimulated controls. The flexibility of ISP stimulation to address focal seizures as well as Lennox-Gastaut syndrome. Efficacy can likely improve with artificial intelligence-based algorithms to optimize and further individualize stimulation parameters. The stimulation target could be reset without further surgical interventions by redesigning the ISP sequence. Machine learning applied to individual and group patient datasets could enhance efficacy, enabling personalized, adaptive treatment for diverse epilepsy conditions.

Other non-invasive strategies, such as EEG-triggered transcranial magnetic stimulation (TMS, ^19–21^) or transcranial direct current stimulation (tDCS, ^22,23^) lack the spatiotemporal precision and immediate ictal engagement provided by ISP stimulation. The observed reduction in seizure duration and modulation of spectral content in our study aligns with the effects of other neuromodulation methods, including direct cortical stimulation and RNS, which alter acute and long-term cortical activity^11,24^. These similarities suggest that ISP stimulation may achieve comparable or better efficacy to invasive neuromodulation techniques, with the added benefit of being minimally invasive. ISP stimulation’s immediate effect in terminating epileptic seizures and the rapid decrease in seizure incidence has advantages versus other neuromodulatory techniques (e.g., VNS, ANT-DBS) with longer build-up durations (i.e., months).

This trial was designed as a proof of concept to prioritize safety and feasibility, but was not powered to establish long-term efficacy. The small cohort size and 10-day study duration with ASM tapering for presurgical evaluation may have influenced seizure incidence. To minimize risks of false-positive detections, semi-automatic stimulus delivery was employed for most seizures, introducing delays in stimulus timing and reducing efficacy in some cases. The limited number of non-stimulated seizures constrained the training of the detection algorithm, underscoring the need for larger datasets to enhance model accuracy and enable fully automated stimulation at seizure onset. The short inpatient duration limit the assessment of long-term disease-modifying effects of ISP stimulation (e.g., sustained reductions in seizure propensity, severity, or duration).

Despite these limitations, our trial demonstrated that ISP stimulation via subgaleal strip electrodes is safe, well tolerated, and technically feasible. The implantation procedure was minimally invasive, with no serious adverse effects, and the electrode configuration enabled precise spatial targeting. The consistent seizure-suppressive effects observed across patients provide compelling feasibility evidence for ISP stimulation as a therapeutic strategy for ASM-resistant epilepsy.

Future studies with longer trial durations, larger cohorts, and advanced stimulation paradigms will be required to establish long-term efficacy, optimize detection algorithms, and explore the broader therapeutic potential of ISP stimulation. With further development, ISP stimulation could represent a transformative, personalized, and minimally invasive treatment for patients with therapy-resistant epilepsy and potentially other neurological disorders.

## Acknowledgments

We sincerely acknowledge the patients for their courage, determination, and willingness to participate in this study, contributing to the advancement of novel therapeutic strategies. We also extend our gratitude to the clinical staff for their dedication and invaluable support in facilitating the successful conduct of this research. This work was supported by the Momentum program II of the Hungarian Academy of Sciences (AB), EFOP 3.6.6-VEKOP-16-2017-00009 (AB), KKP133871/KKP20 (AB), 151490/EXCELLENCE_24 (AB) and 2021-1.1.4-FAST TRACK-2022-00073 (Neunos ZRt) grants of the National Research, Development and Innovation Office, Hungary, the 20391-3/2018/FEKUSTRAT (AB) grant of the Ministry of Human Capacities, Hungary, the EU Horizon 2020 Research and Innovation Program (No. 739593 - HCEMM to AB), Ministry of Innovation and Technology of Hungary grant (TKP2021-EGA-28 to AB), the Hungarian Brain Research Program (grant NAP2022-I-7/2022 to AB).

## Author Contributions

D.F. and A.B. conceived the project.

Z.C., D.F., M.S., L.B., A.P., B.H., M.G., T.F., L.A., T.L., L.H.,

M.HK., N.F., G.S., Á.N., A.Ke., A.S., Z.J., Á.U., A.Ka., L.E.

and A.B. performed clinical duties throughout the study.

M.S., Z.C., M.G., M.HK. and A.B. analyzed the data.

M.S., Z.C., O.D., and A.B. wrote the manuscript.

A.Ka., G.B., O.D., L.E., and A.B. advised the project.

O.D., L.E., and A.B. supervised the project.

## Methods

### Ethical Permission and Protocol

The study was conducted in accordance with the ethical standards set forth in the Declaration of Helsinki. This study was approved by the National Institute of Pharmacy and Nutrition, Hungary (Permission Number: OGYÉI/9674/2021) and the Medical Research Council, Hungary (Permission Number: IV/1920-1/2021/EKU). All participants provided informed written consent after receiving comprehensive information about the study’s objectives, procedures, potential risks, and benefits. Strict clinical protocols were followed to ensure the safety and well-being of the participants throughout the study. All measurements were conducted at the National Institute of Mental Health, Neurology and Neurosurgery (currently the Clinic for Neurosurgery and Neurointervention, Semmelweis University), Budapest, Hungary. The trial is registered at ClinicalTrials.gov (identifier NCT07041619).

### Patient Selection

Patients with frequent focal seizures, with or without bilateral spread, and identifiable ictal EEG features were selected from the patient pool of the Epilepsy Unit of the Institute. Upon consent, their epilepsy histories, previous interictal and ictal EEGs, semiology, neuropsychologic, and neuroimaging data (CT, MRI) were reviewed. Clinical data informed the identification of the most likely seizure onset zones, guiding the surgical and focal stimulus sequence planning.

Fifteen patients were selected, consented, and pre-evaluated; two patients withdrew their consent before implantation. Thirteen patients were implanted and monitored. One session was terminated before stimulating seizures due to cable breakage, which happened during ictal confusion, while twelve patients completed the experimental protocol. The demographic and relevant epileptological data are summarized in Extended Data Table 3. All patients, except for Patient 1 and Patient 15, who were diagnosed with Lennox-Gastaut Syndrome, suffered from intractable focal epilepsy. Etiologies in our patient cohort were bilateral hippocampal sclerosis (n = 1), unilateral hippocampal sclerosis (n = 1), tuberous sclerosis (n = 1), post-NMDAR encephalitis (n = 1), focal cortical dysplasia (n = 2), subependymal heterotopia and polymicrogyria (n = 2), paraventricular heterotopy (n = 1), post-limbic encephalitis (n = 1) and MRI negative (n = 4). Three patients had focal to bilateral tonic-clonic seizures during the study. The small number in our patient cohort does not allow us to draw any conclusion about the efficacy of IPS stimulation based on the seizure type, etiology, or localization.

### Equipment used during the clinical trial

The new modality under investigation is the therapeutic closed-loop ISP stimulation for drug-resistant focal epilepsy. The platform (hereinafter referred to as the Neunos SeizureStop device) is designed to record, detect, and stimulate arising seizure activity. This incorporates four implanted and externalized eight-contact subgaleal electrode array strips (MS08R-IP10X-0JH and MS08R-IP10X-000, Ad-Tech Medical Instrument Corporation, WI, USA), a custom-made tabletop device for both the recording and analysis of EEG activity and closed-loop delivery of ISP stimuli, and the related operating/supporting software. The tabletop device consists of a signal amplifier, a real-time 32-channel EEG processing unit including machine learning-based feature extraction and classification, and stimulation electronics. The device performs real-time seizure detection and attempts to disrupt seizure activity by delivering high-intensity, ultra-short electrical impulses in repeating sequences (i.e., ISP stimulation^16^) to interfere with initiating seizure oscillations. We assume that closed-loop ISP stimulation could offer an effective, minimally-invasive solution for reducing the burden of epilepsy by interfering with the ictal brain activity before behavioral manifestation arises in patients with therapy-resistant epilepsy.

### Study Pipeline

The study pipeline is summarized in Extended Data Figures 2 and 3 and detailed below.

### Preoperative Imaging and Target Planning

The default electrode strip locations were intended to be running in the anteroposterior axis symmetrically 2-3 centimeters from the midline and another pair approximately above the Sylvian fissure. These locations were individualized and adjusted based on the suspected seizure onset zones to ensure maximal spatial coverage of EEG recordings while maintaining proper focusing of stimulated brain volumes of suspected seizure onset zones.

The planning started with an anatomical head-MRI scan to target the electrode implant sites and model the stimulated brain volume. T1 and T2 sequences were obtained. The MRI images were automatically segmented into six tissue/material categories (white matter, gray matter, CSF, skull, scalp, and air) based on the specific intensities of the voxels, using FMRIB Software Library’s (FSL) brain extraction tool and automated segmentation toolbox (FSL, Oxford, UK). Performance and segmentation precision were enhanced by segmenting separately both T1 and T2-weighted sequences of the same head and merging the results. 3D volumetric and cross-sectional reconstructions of the various tissues were generated, visualized, and saved for user supervision purposes.

Next, a finite element model was built from the segmented 3T MRI images^25^. The lead fields of each electrode of the 10/10 system were calculated, and the most optimal linear combination of the calculated lead fields was used to reach the highest relative intensity and focality at the desired target voxel. Next, we aligned the eight-contact strips to the suggested 10/10 locations with consideration for surgical feasibility and also ensured a possible best fit on the suggested 10/10 locations. Finally, the lead fields were recalculated with the defined strip contact coordinates, and an ideal combination and sequence of these lead fields were calculated to maximize the focality of the resulting electrical gradients to the suspected seizure onset zones. This simulation pipeline was aimed to ensure that the ISP stimulation would result in a high current density in relevant brain volumes and a low current density outside of the epileptogenic area, minimizing the side effects. Further adjustments of the strip locations were made intraoperatively as the surgical situation required.

### Implantation and Postoperative Modeling

All surgical implantations were done by an experienced functional neurosurgeon under general anesthesia. The strips were inserted using posterior and if needed additional anterior incisions for each strip. The strips were pushed from posterior incision, or pulled with a thread temporarily secured at the tip of the strip from the anterior incision. Strip locations were confirmed by neuronavigation and, in some cases, a hybrid method with additional C-arm control when deemed necessary^26^. The final strip locations were verified by a post-operative non-contrast CT scan and if the deviation from the planned locations required, the ISP sequences were recalculated to ensure the precision of focal stimulation.

### Data acquisition

The EEG was acquired by the Neunos SeizureStop device (by Neunos ZRt, Szeged, Hungary) and is saved in a proprietary format. The sampling rate was 500 Hz, and the signal was band-pass filtered between 1 Hz and 250 Hz. The channels were referenced to one of the strip contacts, which was chosen to be the most distant location from the suspected seizure onset zone, or to a common average of the electrodes. Besides the acquired EEG signals, the device also calculated and stored 17 features of each of the 33 EEG channels (32 physical channels and a virtual channel derived from the weighted linear combination of physical channels’ signals) using a sliding window (1000 ms window size, 900 ms overlap). These time- and frequency-domain feature values numerically characterize the properties of the EEG signal in the time-frequency domain (e.g., dominant frequency, amplitude, skewness, coherence, etc.), within the one-second-long windows, updated at 10 Hz.

### Baseline Recordings and Seizure Types

After obtaining the post-operative CT scan, the patients were placed in the monitoring room in the Epilepsy Monitoring Unit of the hospital. The implanted electrode strips were connected to the Neunos SeizureStop device and also routed to a clinical video-EEG recording system for backup and clinical diagnostic purposes. Except for periodic powerline artifacts, we managed to obtain state-of-the-art quality EEG recordings throughout the experiment. The Epilepsy Unit’s medical staff carefully adjusted the dosage of anti-epileptic drugs when they determined it was essential. This controlled reduction aimed to facilitate the recording of an adequate number of seizure events for post-hoc analysis. Baseline EEG and seizure activity were monitored and recorded uninterruptedly while an experimenter continuously supervised the system. After collecting 3-4 days of baseline data, including awake EEG, sleep EEG, and artifacts, as well as observed habitual seizures, we proceeded to train the detector algorithm using the recorded and annotated EEG epochs.

### Stimulation Intensity Tolerance Titration

We thoroughly evaluated the individual stimulation tolerance thresholds of the patient to minimize discomfort during the ISP stimulation in the event of seizures. We employed the pre-modeled ISP sequences, which were later used for ictal therapeutic stimulation. The patients’ thresholds were defined by using a Visual Analog Scale (VAS) method while gradually increasing the stimulation amplitude. We recorded all VAS values reported by the patients and recorded all sensations that were verbally reported after each stimulus. We also interviewed patients after each stimulated seizure to record their subjective perceptions and sensations.

### Seizure Stimulation

After we recorded habitual seizures and trained the seizure detector on those, we started the seizure stimulation protocol of the experiment. We employed charge-balanced ISP stimulation patterns (Extended Data Figure 1). The rotating dipole switching sequences were wrapped in a 50 ms long half-sinusoid waveforms^18^. The half sine envelope ensured that the maximum amplitude within a sequence was ramped up, thus decreasing the peripheral side effects. The half sine shape of the stimuli also resembles spontaneously occurring oscillating brain activity driving the synchronization of cortical neurons at lower frequencies. Thus, we hypothesized that sinusoidal rise and fall of intensity are better suited for disrupting pathological synchrony in the epileptic cortex. The half sines were repeated one to three times with a rate matched to the dominant frequency of the initiating seizure as defined by the control records. Once a seizure was detected by the algorithm, visual and auditory alerts were sent to investigators for confirmation of behavioral and EEG signs of a seizure before triggering preprogrammed ISP stimulus delivery (semi-automatic delivery). Although the investigator’s confirmation in semi-automatic stimulus delivery mode introduced a significant delay in delivering the stimuli, it was necessary for patient safety in this pilot experiment. An automatic mode was used for highly reliable detections in select patients under close human expert supervision. Fully manual triggering of interventions was also employed when EEG and semiology signs indicated a seizure onset, but the detector failed or delayed alerting the investigators.

### Explantation, Emission, and Follow-up

Once measurements were concluded, the electrode strips were explanted under general or local anesthesia. Patients were followed up by scheduled surgical and epileptological visits, where control of the surgical wounds and changes in seizure frequency were assessed.

### EEG Annotation

To compare control and stimulated seizures, we carefully annotated seizures post hoc, according to clinical conventions. Annotations were based on the recorded EEG landmarks supported by the video recordings of the clinical correlates. Epileptiform seizure patterns (ESPs) were visually identified by experienced EEG readers, aided by peri-ictal spectrograms. The onset of ESPs was marked at the first epoch, after which the EEG recording was changed from interictal background activity and was followed by a paroxysmal discharge of seizure features on any channel or at abrupt stereotypical ictal behavioral change. The end of seizures was marked when the EEG ictal activity ceased. If a stimulation overlapped with the end of seizures and the stimulation artifact interfered with the ESP visibility, we marked the end where the stimulation artifact was no longer masking the EEG signal. Due to the presence of visible stimulus artifacts, blinded annotations were not possible; however, two experienced EEG readers annotated the recordings independently, and a consensus was reached when necessary regarding the seizure lengths.

### Post Hoc Signal Processing and Data Analysis

All analyses described in this section were performed with custom-written Python and Matlab scripts (version 3.12 and 2024b, respectively). Two-sample Kolmogorov-Smirnov test was used for comparison with Bonferroni correction for multiple comparisons, unless otherwise noted. One, two and three asterisks denote significance levels of < 0.05, < 0.01, and < 0.001, respectively.

The spectral characteristics of control and simulated seizures were assessed using a single channel with the highest seizure-related SNR. Spectrograms were computed using continuous wavelet transformation with a Morlet wavelet (cmor1.5-1.0 in pywt), and the resulting power values were smoothed in time with a Gaussian filter (σ = 2). Spectral density estimation of the seizure epochs was computed using Welch’s method implemented in scipy’s signal processing package (with the parameter nperseg = 1024). Power spectral densities (PSDs) were maximum-normalized and averaged. Statistical comparison between the mean normalized PSDs were performed with a two-sample Kolmogorov-Smirnov test individually in each of the four frequency bands (delta (0.5-4 Hz), theta (4-8 Hz), alpha (8-13 Hz), and beta (13-30 Hz)). Outliers were removed as the 99^th^ percentile of data.

For analyses in relation with seizure durations, stimulated seizures of the treatment period were compared to the seizures of the baseline period. Cases involving incomplete stimulation resulting from technical errors were omitted from the analyses. To analyze the effect of stimulation delay on seizure termination efficacy, patients with more than five stimulated seizures were included. Intra- and interpatient variability of seizure durations were compensated by normalizing each stimulated seizure’s length to the mean duration of the non-stimulated seizures of the given patient. The delay of the stimulation was normalized the same way.

In the seizure incidence analysis, we included patients with statistically robust seizure incidences (daily fluctuation < 4 SD), long enough stimulated periods (> 48h), continuous recordings (total non-recorded periods < 5% of total experiment time) and high-coverage stimulation of ictal events (> 95% of seizures stimulated during the stimulation period). Baseline seizure incidence was calculated from the period between the first postoperative seizure (often absent on days 1– 2 due to residual effects of anesthesia) and the start of ictal stimulation. The treated period incidence was then evaluated cumulatively, starting with the entire stimulated period from the onset of stimulation until hospital discharge, and then recalculated multiple times while progressively shifting the start point forward in time (e.g., beginning 24 hours after stimulation onset, then 48 hours, and so on). All electroclinical seizures occurring within the specified periods, encompassing both stimulated and non-stimulated events, were incorporated into the analyses. A sliding window spanning 12 hours and shifted by 4 hours in each step was used within each segment to generate surrogate data for statistical comparison of seizure incidence rates.

## Data Availability

The raw data generated and analyzed in this study are proprietary to Semmelweis University, University of Szeged and Neunos ZRt and include sensitive patient health information protected under the EU General Data Protection Regulation (GDPR). Due to these intellectual property restrictions and privacy considerations, the raw data cannot be made publicly available. De-identified summary data supporting the findings are provided in the article and its Supplementary Information. Additional data may be available from the corresponding author upon reasonable request and subject to institutional and legal approval.

## Code Availability

All custom codes are freely available from the corresponding author on reasonable request.

## Extended Data

**Extended Data Fig. 1.**
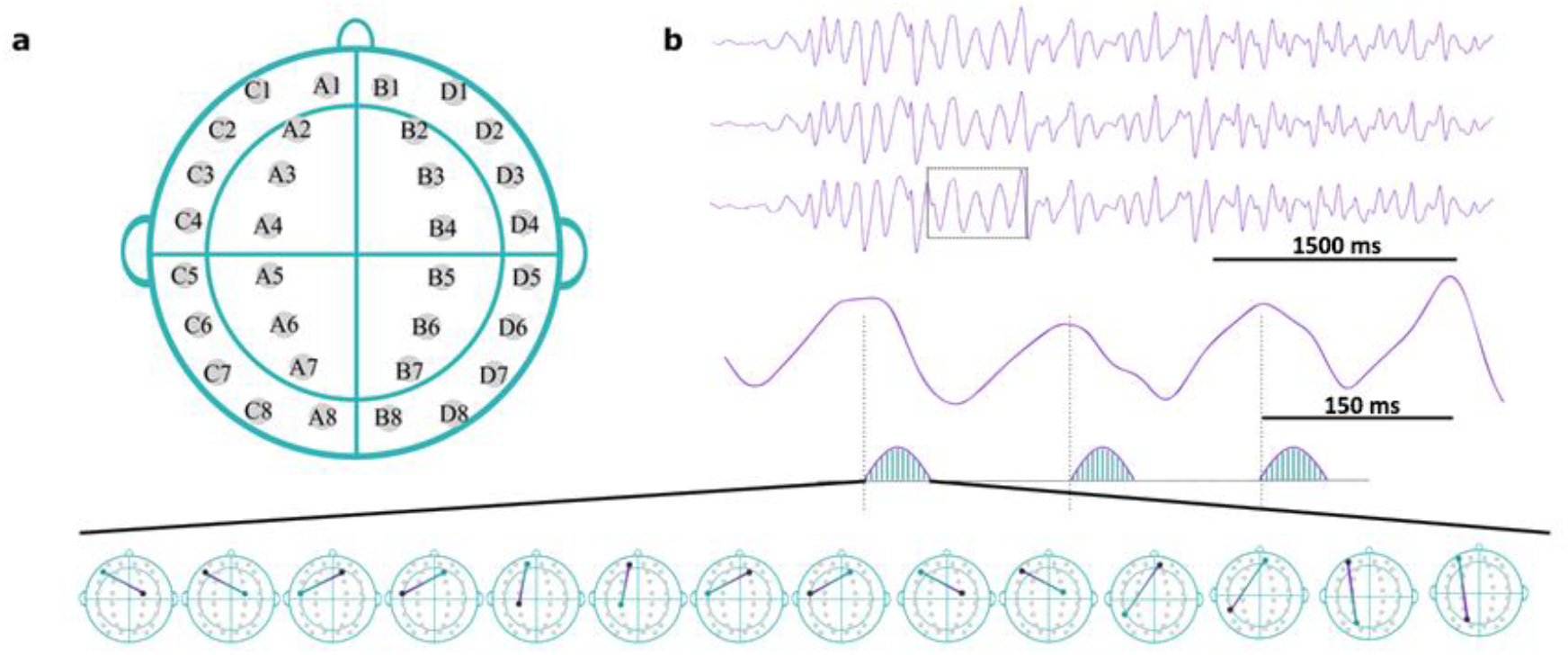
ISP stimulation’s mode of effect. **a**, Subgaleal strip electrode configuration. Four electrode strips (C, A, B, D, left temporal, left parasagittal, right parasagittal, and right temporal, respectively), each having eight contact sites, were implanted in each patient. **b**, Schematic representation highlighting the temporal alignment of ISP stimulation with the ongoing seizure activity. The figure depicts one ‘dose’ of ISP stimulation consisting of three repeats of 50-millisecond long trains of 100 µs wide ISP pulses. The delay between the three repeats is aligned with the dominant frequency of the seizure to consistently target the same phase of the seizure oscillations. The 50-millisecond half-sine envelope modulates the stimulus intensity smoothly to minimize adverse sensory effects. Each half-sine envelope comprises a sequence of individual pulses with changing amplitudes, arranged across different electrode pairs. This precise temporal and spatial modulation are designed to maximize stimulation efficacy at the epileptic focus while minimizing discomfort and collateral effects on non-target areas.

**Extended Data Fig. 2.**
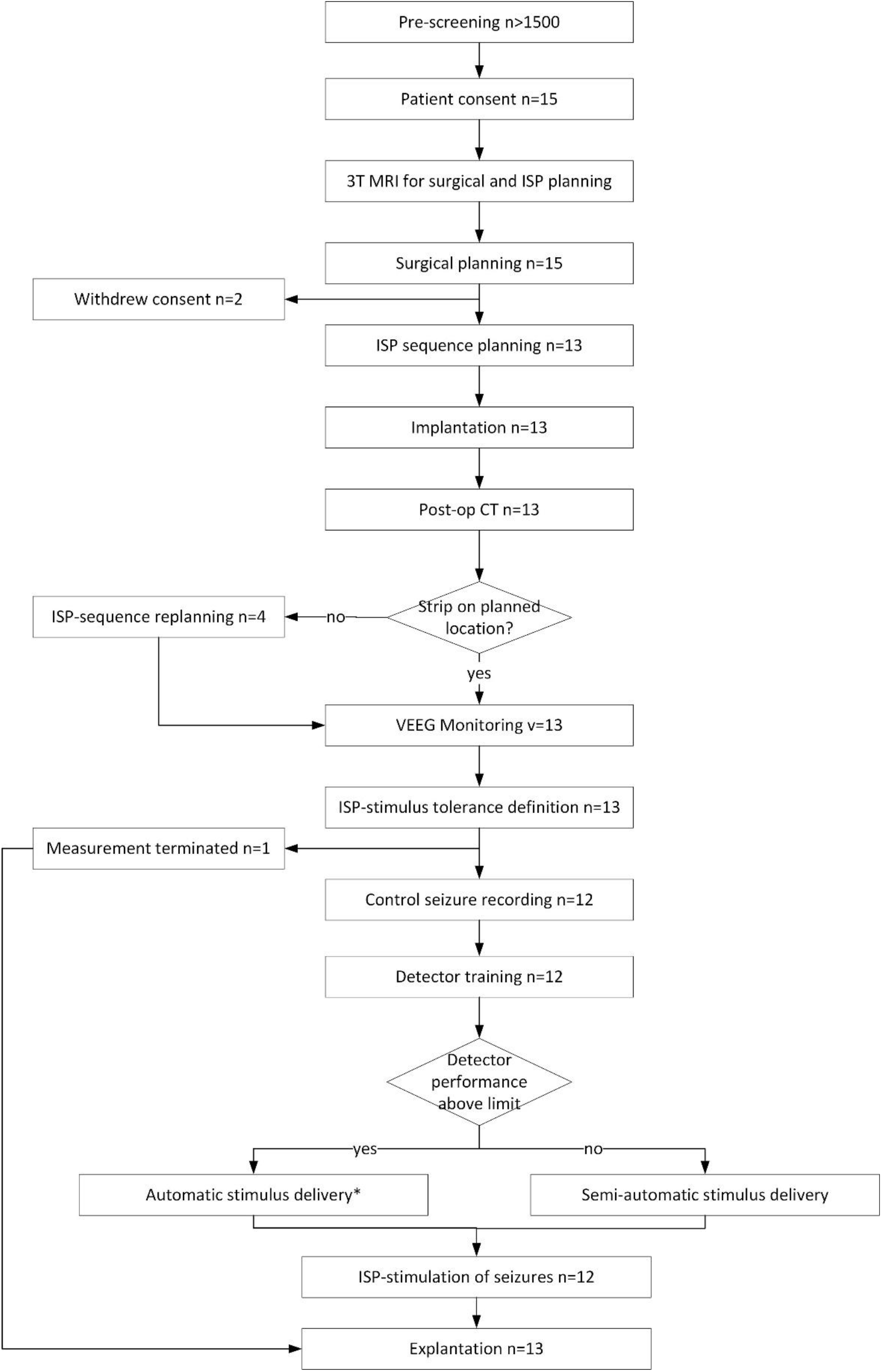
Flowchart of the study pipeline.

**Extended Data Fig. 3.**
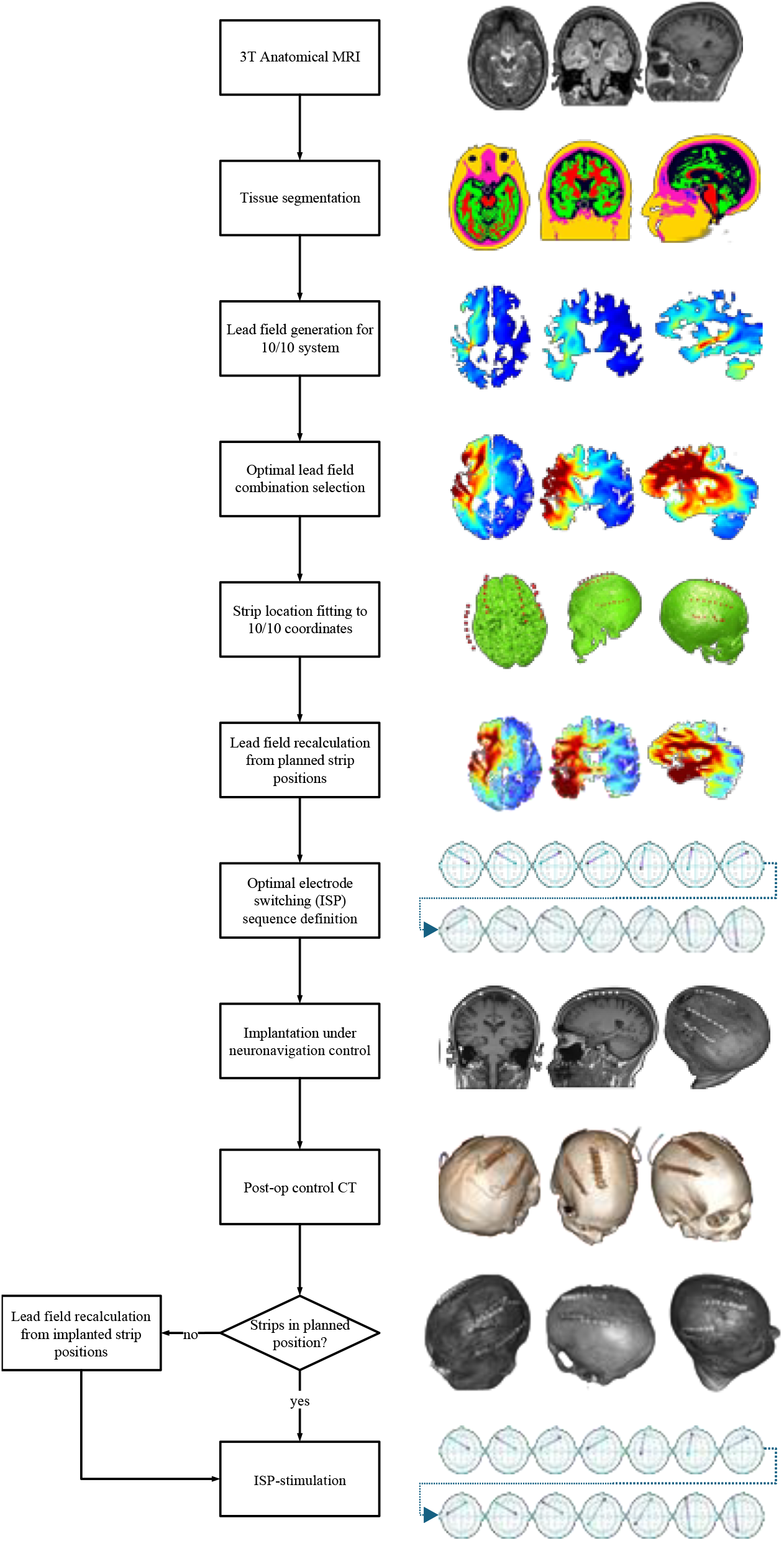
The seizure onset zone targeting process used for ISP stimulation optimization and the corresponding representative images from Patient #2.

**Extended Data Fig. 4.**
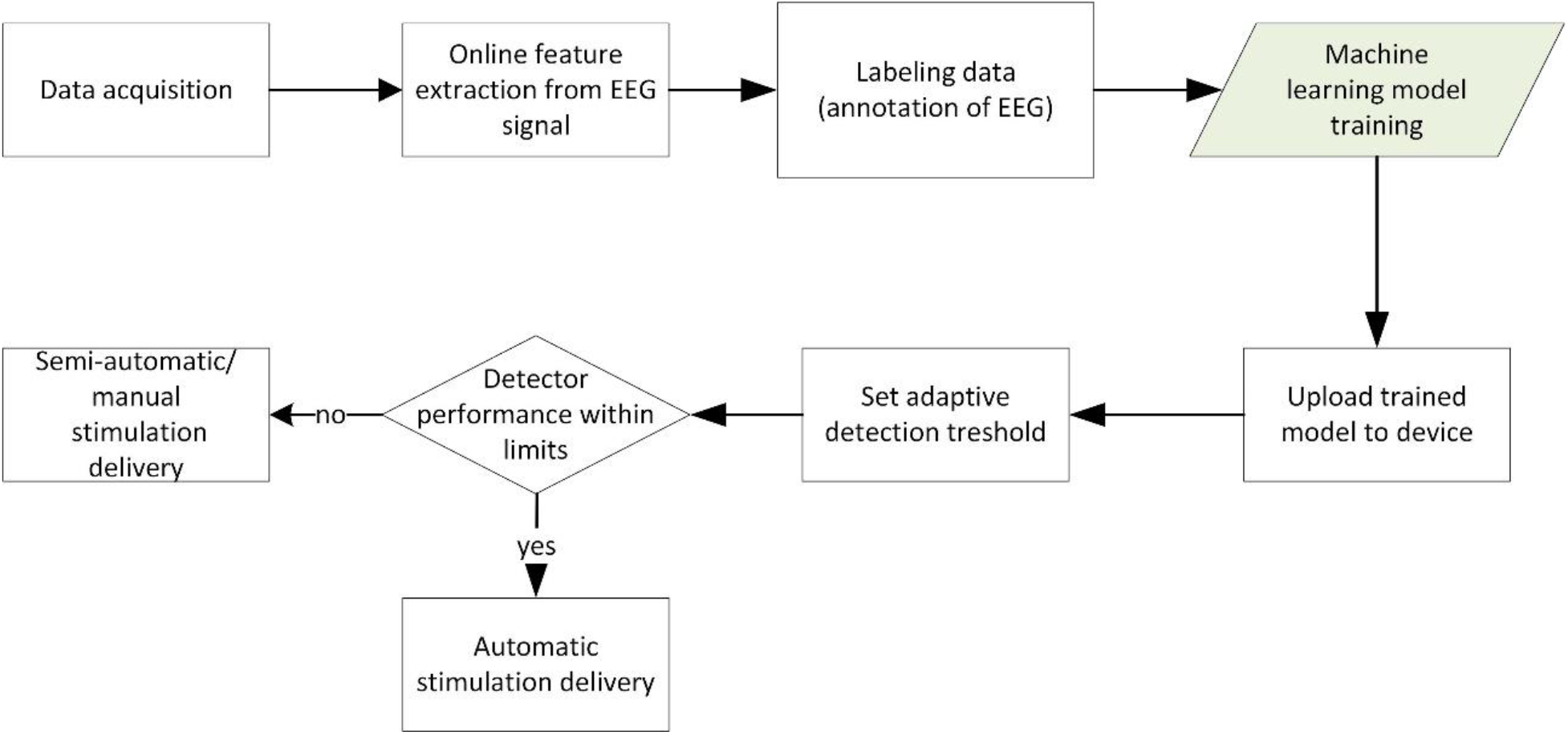
Flowchart of seizure detector training for real-time EEG-based seizure detection.

**Extended Data Fig. 5.**
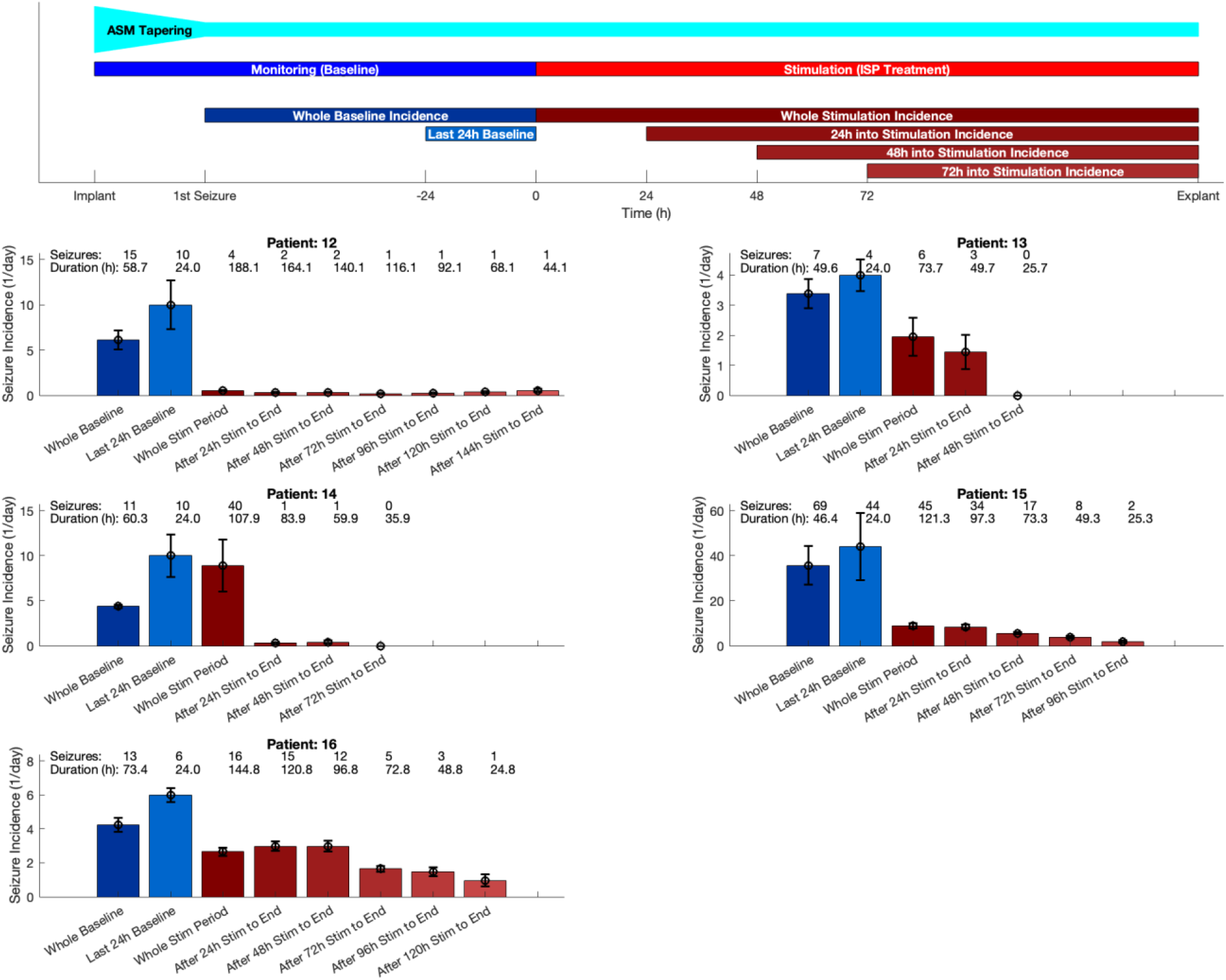
Per-patient seizure incidence and analysis protocol. The top panel illustrates the experimental timeline for each patient, including ASM tapering, monitoring, and ISP stimulation periods, with breakdown of analysis windows. The baseline seizure incidence was calculated from the appearance of the first seizure to the initiation of ictal ISP stimulation. The stimulated periods’ incidences were evaluated cumulatively from different starting points (whole stimulation period, 24 h after stimulation onset, 48 h after onset, etc.) to capture the progressive therapeutic effect. The lower panels show per-patient seizure incidences during the baseline and stimulation periods (mean ± s.e.m.). Blue bars represent baseline values, red bars represent stimulation period values, with lighter shades indicating progressively later analysis start points, and consequently longer pre-analysis treatment periods. Numbers above each bar denote total seizure counts during the respective period, and the duration of the analyzed periods (in hours).

**Extended Data Fig. 6.**
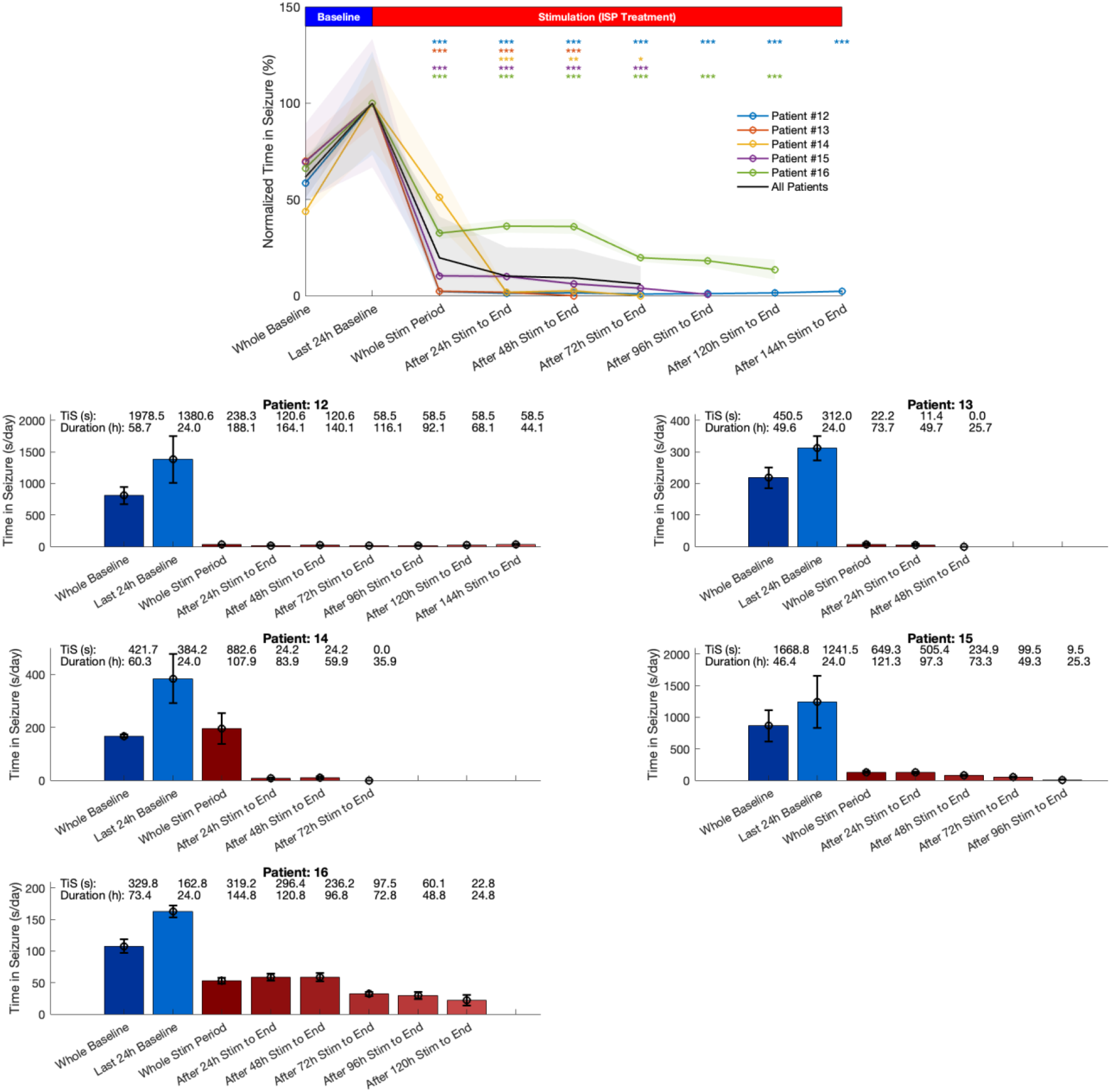
Analysis of the total time spent in seizure. The top panel illustrates the summary of the times spent in seizures for each patient, normalized to the last 24 hours of the baseline period. The lower panels show the non-normalized per-patient time in seizure data during the baseline and stimulation periods (mean ± s.e.m). Numbers above each bar denote total time spent in seizure during the respective period (in seconds), and the duration of the analyzed periods (in hours). Analysis methodology and conventions are otherwise the same as on Figure 4b and Extended Data Fig 5.

**Extended Data Table 1.** Summary of adverse events by main categories and subcategories. The top table shows cumulative days affected, number of events, number of patients, and severity, while the bottom matrix details the distribution of events per patient across categories. Most AEs were mild, transient, and related to the surgical implantation procedure. Importantly, all stimulation-related events were mild, and no serious adverse event was attributable to the therapy. The two grade 3 events (psychosis due to antiseizure medication tapering and fever due to urinary infection) were unrelated to ISP stimulation. The “Other study related” category includes AEs that were linked to the broader study context (e.g., AED tapering, antibiotic side effects, protocol-related clusters) but not to the surgery or stimulation itself. All adverse events fully resolved, either spontaneously or with appropriate therapy.

| Main Category | Subcategory | Total Days affected | Number of Events | Number of Patients affected | Severity (Mean ± SD, [Range]) |
| --- | --- | --- | --- | --- | --- |
| Surgery / Anesthesia | Anesthesia-related systemic | 5 / 116 | 5 | 4 / 12 | 2.00 ± 0.00 [2–2] |
| | Incision/wound & healing | 11 / 116 | 12 | 9 / 12 | 1.30 ± 0.48 [1–2] |
| | Hematoma/edema | 13 / 116 | 4 | 4 / 12 | 1.00 ± 0.00 [1–1] |
| | Other peri-surgical | 19 / 116 | 17 | 11 / 12 | 1.33 ± 0.50 [1–2] |
| Therapy Delivery | Impedance measurement | 9 / 116 | 2 | 2 / 12 | 1.00 ± 0.00 [1–1] |
| | Stimulation sensations | 10 / 116 | 8 | 6 / 12 | 1.29 ± 0.49 [1–2] |
| | Other device-related | 11 / 116 | 7 | 6 / 12 | 1.33 ± 0.58 [1–2] |
| Other Study Related | Medication/AED tapering | 1 / 116 | 1 | 1 / 12 | 3.00 ± 0.00 [3–3] |
| | Protocol-related | 6 / 116 | 6 | 4 / 12 | 1.67 ± 0.52 [1–2] |
| Other | Infections/inflammatory | 6 / 116 | 1 | 2 / 12 | 2.00 ± 1.41 [1–3] |
| | Respiratory/GI | 3 / 116 | 3 | 2 / 12 | 1.33 ± 0.58 [1–2] |
| | Sleep-related | 2 / 116 | 2 | 2 / 12 | 1.00 ± 0.00 [1–1] |
| | Miscellaneous | 18 / 116 | 10 | 8 / 12 | 1.00 ± 0.00 [1–1] |

**Extended Data Table 1 | Summary of adverse events by main categories and subcategories.**
|  |  | Patient ID |  |  |  |  |  |  |  |  |  |  |  |  |
| --- | --- | --- | --- | --- | --- | --- | --- | --- | --- | --- | --- | --- | --- | --- |
| Main Category | Subcategory | 1 | 2 | 3 | 4 | 5 | 7 | 9 | 10 | 12 | 13 | 14 | 15 | 16 |
| Surgery / Anesthesia | Anesthesia-related systemic | 0 | 2 | 2 | 0 | 0 | 0 | 0 | 1 | 0 | 0 | 0 | 0 | 0 |
|  | Incision/wound & healing | 1 | 0 | 2 | 0 | 1 | 2 | 1 | 1 | 1 | 0 | 0 | 2 | 1 |
|  | Hematoma/edema | 1 | 0 | 0 | 0 | 0 | 0 | 0 | 0 | 1 | 0 | 1 | 1 | 0 |
|  | Other peri-surgical | 0 | 4 | 2 | 2 | 2 | 2 | 0 | 2 | 0 | 0 | 2 | 1 | 0 |
| Therapy Delivery | Impedance measurement | 0 | 0 | 0 | 0 | 0 | 0 | 0 | 0 | 0 | 1 | 1 | 0 | 0 |
|  | Stimulation sensations | 0 | 1 | 1 | 1 | 0 | 0 | 0 | 0 | 1 | 0 | 1 | 2 | 1 |
|  | Other device-related | 1 | 0 | 0 | 0 | 0 | 3 | 1 | 1 | 1 | 0 | 0 | 0 | 0 |
| Other Study Related | Medication / AED tapering | 0 | 0 | 0 | 0 | 0 | 0 | 0 | 0 | 1 | 0 | 0 | 0 | 0 |
|  | Protocol-related | 0 | 1 | 0 | 0 | 0 | 0 | 0 | 0 | 0 | 1 | 1 | 3 | 0 |
| Other | Infections/ inflammatory | 0 | 0 | 0 | 0 | 0 | 0 | 0 | 0 | 0 | 0 | 0 | 1 | 0 |
|  | Respiratory/GI | 0 | 0 | 1 | 0 | 0 | 0 | 0 | 2 | 0 | 0 | 0 | 0 | 0 |
|  | Sleep-related | 0 | 0 | 0 | 0 | 0 | 0 | 0 | 0 | 1 | 1 | 0 | 0 | 0 |
|  | Miscellaneous | 0 | 0 | 2 | 0 | 0 | 0 | 1 | 1 | 2 | 3 | 0 | 1 | 0 |
| Total No. of Events |  | 3 | 8 | 10 | 3 | 3 | 7 | 3 | 8 | 8 | 6 | 6 | 11 | 2 |

**Extended Data Table 2.** Descriptive Statistics and Statistical Comparison of ISP Stimulation Effects on Seizure Duration. For Patient #7, ^†^ and ^×^ denote left temporal and bilateral ISP stimulation paradigms, respectively. P-values for Patient #7 and #10 are not reported due to the very low number of observations.

| Patient # | Baseline Seizures |  | ISP Stimulated Seizures |  | Seizure Shortening | Mean Stimulation Delay (s) | Delay-compensated Seizure Shortening | P-value |
| --- | --- | --- | --- | --- | --- | --- | --- | --- |
| | Mean ± SD (s) | N | Mean ± SD (s) | N | | | | |
| 1 | 4.66 ± 3.17 | 115 | 1.40 ± 0.81 | 63 | -70% | 1.35 | -98% | < 0.001 |
| 2 | 128.80 ± 56.53 | 5 | 43.88 ± 10.98 | 4 | -66% | 29.38 | -85% | 0.007 |
| 3 | 32.25 ± 0.02 | 2 | 33.86 ± 8.65 | 13 | 5% | 9.04 | 7% | 0.514 |
| 7 | 110.00 ± 0.00 | 1 | 73.10 ± 3.52<sup>†</sup> | 2 | -34% | 20.71 | -41% | N/A |
| | | | 34.35 ± 4.51<sup>×</sup> | 2 | -69% | | -85% | N/A |
| 9 | 80.98 ± 107.38 | 23 | 24.74 ± 38.74 | 28 | -69% | 10.87 | -80% | < 0.001 |
| 10 | 70.05 ± 6.66 | 2 | 56.41 ± 0.80 | 2 | -19% | 16.89 | -26% | N/A |
| 12 | 131.90 ± 58.63 | 15 | 59.56 ± 5.10 | 4 | -55% | 34.50 | -74% | < 0.001 |
| 13 | 64.35 ± 29.34 | 7 | 3.70 ± 1.72 | 6 | -94% | 1.64 | -97% | 0.001 |
| 14 | 36.94 ± 6.94 | 12 | 22.08 ± 5.99 | 39 | -40% | 5.07 | -47% | < 0.001 |
| 15 | 24.11 ± 14.49 | 71 | 14.10 ± 8.74 | 43 | -41% | 3.75 | -49% | < 0.001 |
| 16 | 23.57 ± 4.83 | 19 | 13.53 ± 7.94 | 18 | -43% | 2.05 | -47% | < 0.001 |

**Extended Data Table 3.**
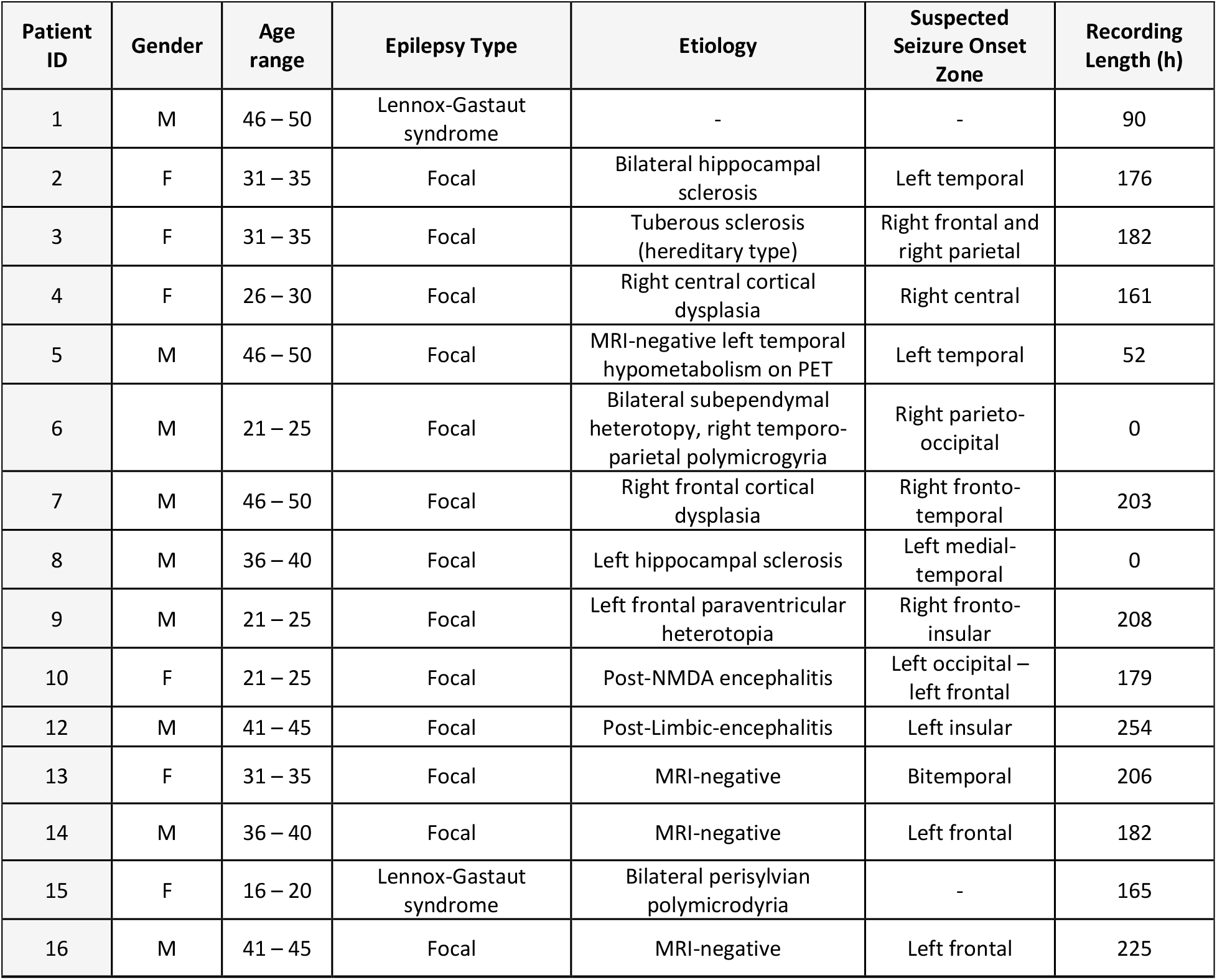
Demographic and epileptology details and total video-EEG recordings of the patients.

**Extended Data Video. 1.**
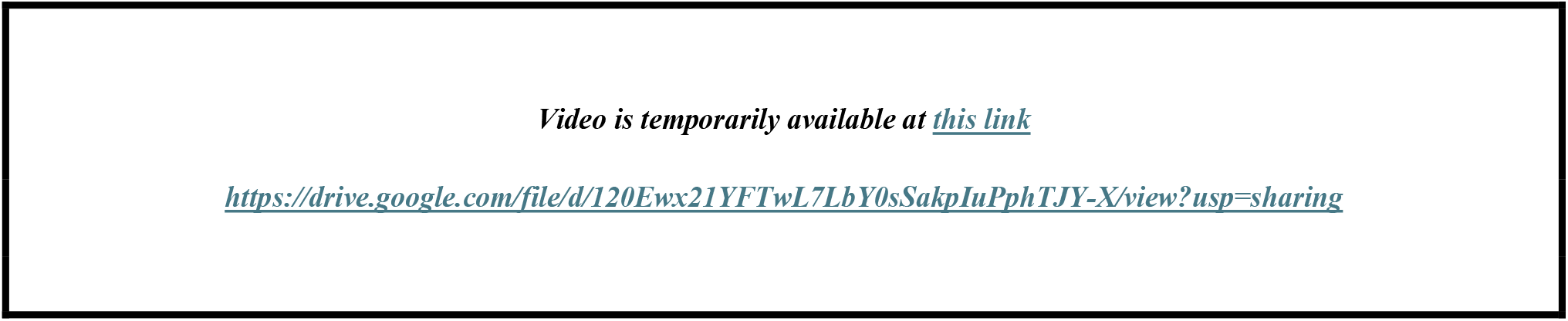
Example video EEG recordings. The video shows two seizures of Patient #2. The non-stimulated control seizure and a stimulated seizure are shown on the upper and lower panel, respectively. The left panes show the 32 EEG channels, color coded by strips as blue, yellow, red and green, and the automated seizure detector’s output (black trace). The right panes show the corresponding video signals synced to the blue, thick vertical line in the middle of the EEG panes. The lower seizure was stimulated twice during the focal phase of the seizure, marked with a red vertical arrow on the recording. Note the abrupt change in the EEG pattern after stimulation and the lack of generalization as opposed to the unstimulated control seizure on the top. (FIAS: focal impaired awareness seizure; FBTCS: focal to bilateral tonic clonic seizure)

